# Deployment of a digital twin using the coupled momentum method for fluid-structure interaction: a case study for aortic aneurysm

**DOI:** 10.1101/2024.11.07.24316881

**Authors:** Giacomo Creazzo, Guido Nannini, Simone Saitta, Davide Astori, Mario Gaudino, Leonard N. Girardi, Jonathan W. Weinsaft, Alberto Redaelli

## Abstract

**Introduction:** Dacron graft replacement is the standard therapy for ascending aorta aneurysm, involving the insertion of a prosthesis with lower compliance than native tissue, which can alter downstream hemodynamics and lead to adverse remodeling. Digital human twins (DHT), based on in-silico models, have the potential to predict biomarkers of adverse outcome and aid in designing optimal treatments tailored to the individual patient.

**Objective:** We propose a pipeline for deploying a digital human twin of the thoracic aorta to explore alternative solutions to traditional Dacron grafting, utilizing more compliant prostheses for reconstructing the ascending aorta.

**Methods:** We propose a DHT based on fluid-structure interaction (FSI) analysis of the thoracic aorta. We create 3 models of the patient, representing: i) the pre-operative baseline, ii) the post-operative with Dacron graft, and iii) a virtual post-operative using a compliant fibrous prosthesis. 3D geometry of the thoracic aorta for a patient with a congenital aneurysm, before and after the surgery, were reconstructed from magnetic resonance imaging (MRI). As inlet boundary condition (BC), we assigned a time-varying 3D velocity profile extrapolated from 4D flow MRI. For the outlet BCs, we coupled 0D Winkler models, tuned to match the flow rate measured in the descending aorta from 4D flow. The aortic wall and the prosthetic graft were modeled as hyperelastic materials using the Holzapfel-Gasser constitutive model and tuned to patients distensibility. FSI analysis was run for two cardiac cycles.

**Results:** Results were validated against 4D flow data. Quantitative comparison of outflows between FSI and 4D flow yielded relative squared errors of 5.28% and 0.33% for models (i) and (ii), respectively. Wall shear stress (WSS) and strain increased in both post-surgical scenarios (ii) and (iii) compared to (i), with a lower increase observed in the virtual scenario (iii) (p¡0.001). However, the difference between scenarios (iii) and (ii) remained moderate on average (e.g., 0.6 Pa for WSS).

**Conclusions:** FSI analysis enables the deployment of reliable thoracic aorta DHTs to predict the impact of prostheses with different distensibility. Results indicate moderate yet promising benefits of more compliant fibrous devices on distal hemodynamics.

**Graphical Abstract:** 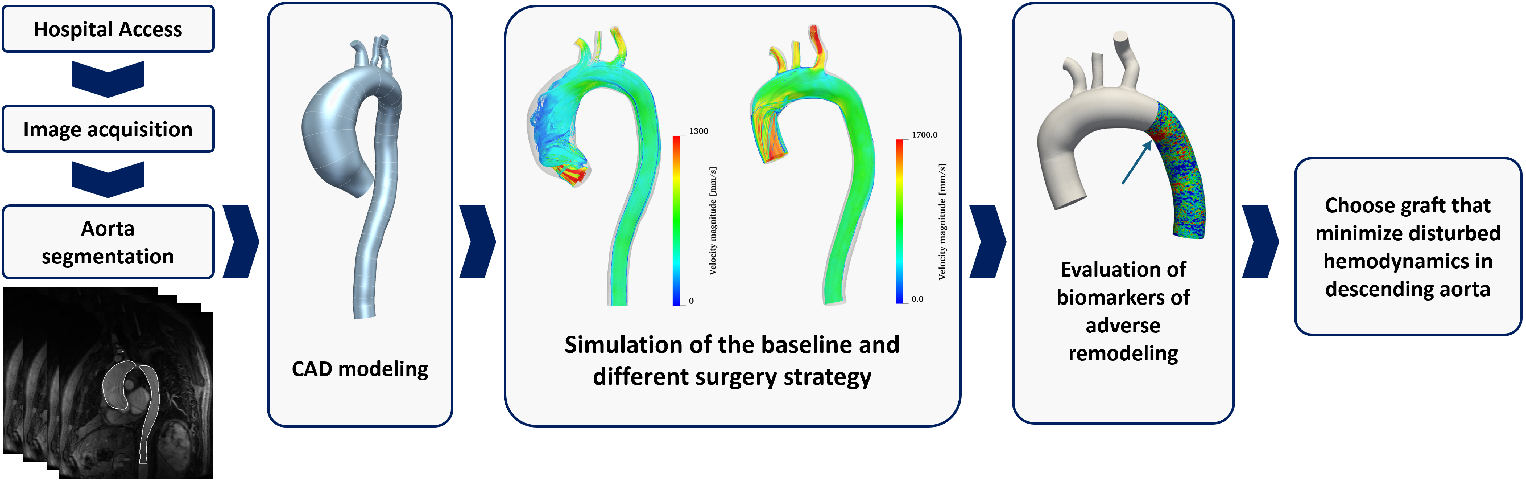

**Highlights:**

- The coupled momentum method for fluid-structure interaction analysis can be used to deploy a high fidelity digital twin of the aorta
- Dacron graft induces an altered hemodynamic state in the descending aorta after ascending aorta correction.
- Electrowriting can be used to fabricate more compliant grafts that mitigate the adverse effect of Dacron in the descending aorta.
- The stress induced by a 2-folds more compliant (with respect to Dacron) graft is significantly lower
- Despite the difference being relevant, it is moderate (1 Pa).

## 1. Introduction

Ascending thoracic aortic aneurysm (ATAA) is one of the most common aortic diseases, with an incidence of 5.3 per 100,000 individuals per year. It involves the localized dilation of the vessel due to vascular wall weakening, which may lead to rupture and sudden death [1]. Prosthetic graft implantation is a safe and beneficial therapy, that consists in the removal of the dilated aorta and reconstruction using *Dacron* [2]; however, ATAA reconstruction leads to permanent alterations in the aortic biomechanics and hemodynamics, exposing patients to the risk of adverse remodeling in the downstream native aorta [3]. The underlying mechanisms for these occurrences remain uncertain, but one plausible explanation is the interaction between the grafted portion and the native aortic tissue. Indeed, the *Dacron* vascular prosthesis is much stiffer than the native aortic wall, resulting in the absence of a compliant reservoir to absorb the kinetic energy of systolic ejection in the ATAA tract and leading to increased flow velocity [4]. Moreover, the surgery reduces the ascending aortic caliber, which further amplifying the delivery of kinetic energy downstream [5]. Altered hemodynamics contributes to internal elastic lamina fragmentation and vessel tortuosity, both of which are associated with the risk of dissection and create a potential nidus for adverse remodeling [3]. To address these issues, research has focused on alternative graft fabrication techniques and materials. Solution electrowriting is an innovative technique that can be used to produce tubular scaffolds with tunable micro-architecture, as well as customizable physical, chemical, and mechanical properties, enabling them to mimic the compliance of physiological vessels [6]. However, a knowledge gap remains regarding whether graft-induced alterations in aortic biomechanics vary depending on the prosthesis length and mechanical properties. Thus, the analysis of flow parameters such as velocity and wall shear stress (WSS) may serve as metrics for assessing the impact of the graft on aortic biomechanics during clinical follow-up [7]. 4D phase-contrast magnetic resonance imaging (PC-MRI), or 4D flow, allows for the *in vivo* measurement of local blood velocity, providing a non-invasive, comprehensive understanding of blood flow dynamics in three dimensions over time. It enables the assessment of complex hemodynamic phenomena, including vortex formation and turbulence. Despite its potential, the limited spatial and temporal resolutions of 4D flow impede the precise quantification of parameters that require velocity space- or time-derivatives (e.g., WSS) [8]. To overcome these limitations, patient-tailored *in silico* approaches based on fluid-structure interaction (FSI) and computational fluid dynamics (CFD) have been widely used, combined with *in vivo* patient-specific data, to create virtual high-fidelity replicas of real-world patients, often referred to as *digital human twins* (DHTs) [13, 14, 15, 16, 17], and investigating in detail the effects of aortic grafting [18, 19, 5]. DHTs have garnered increasing interest as tools to support clinical decision-making, due to their ability to systematically predict the effects of specific surgeries by identifying key indices considered pivotal for predicting post-operative major adverse events [20]. Thus, *in silico* modeling algorithms, such as CFD, *2-way* FSI [11] and coupled momentum method for FSI (CMM-FSI) [12], set the basis for the deployment of an effective physics-informed DHT. Furthermore, DHT models provide high spatial and temporal resolution, offering unique insights into the complex interplay between blood flow and the vascular wall, as well as the ability to test the impact of actual or simulated alterations in material properties on factors such as WSS or mechanisms of adverse remodeling [5].

Rocatello *et al*. [21] presented an aortic DHT for transcatheter aortic valve implantation planning, based on finite element method analysis on 62 patients. The DHT was used to predict adverse outcome of the surgery, such as paravalvular leakage, when implanting different prosthesis, and was validated against postoperative follow-up. In their work, Groth *et al*. [22] combined reduced order models (ROM) with mesh morphing techniques to develop a DHT for ATAA and study the effects of bulge shape progression on the flow field. This innovative approach ensures real-time, patient-specific predictions of hemodynamic changes under various aneurysm shapes, providing rapid insights to support personalized surgical planning for ATAA. Salica *et al*.[23] proposed a DHT for mitral valve annuloplasty, to simulate the implanting of annuloplasty rings of different sizes and predict the corresponding tissue strains, and suture forces, which identify areas at risk for dehiscence.

The development of a DHT requires the following steps: *i*) Reconstruct the anatomy of the district of interest by segmenting medical images; *ii*) Spatially discretize the geometry to create a computational mesh; *iii*) Identify an accurate set of boundary conditions (BCs) for reliable clinical predictions and describe the material properties using an appropriate constitutive law. A common BCs choice is to couple a 4D flow velocity field at the inlet with three-element Windkessel models (3WKMs) at the outlets. [9, 10]; *iv*) To ensure the reliability of the model, validation against *in vivo* measured data must be achieved; *v*) The DHT can finally be used to infer the quantities of interest. In our work, we focus on alternatives to the commonly used *Dacron* prosthesis for ATAA correction, aiming to utilize CMM-FSI modeling to implement a DHT, enabling a comprehensive analysis of biomechanical indeces that are altered by ATAA correction, and are strong indicators of adverse remodeling [3]. Thus, we analyze one patient as a *proof-of-concept* and we propose a pipeline for implementing a robust DHT that can be used to predict surgical outcomes and explore tailored clinical solutions. We developed three aortic models representing: *i*) The baseline; *ii*) The actual post-surgery scenario, with the ATAA reconstructed using a *Dacron* graft (DG). Model validation against clinical data was performed to ensure the reliability and predictive capability of the DHT. *iii*) Finally, a virtual post-surgery configuration that uses a compliant electrowritten graft (EW) was implemented to investigate the influence of stiffness mismatch on distal hemodynamics.

## 2. Materials and methods

### 2.1. Dataset

Magnetic resonance (MRI) was acquired on one subject suffering from congenital ATAA, who underwent graft implantation at New York-Presbyterian Hospital (New York, NY, USA). Contrasted enhanced MRI angiography was acquired with the following setup: voxel size = 0.752 × 0.752 × 2 mm, field of view (FOV) = 380 mm, flip angle = 30°, repetition time (T_R_) = 4 ms, echo time (T_E_) = 1.56 ms. Subsequently, PC-MRI was acquired with sagittal-oblique orientation, spatial resolution was set to an isotropic value of 1.8 × 1.8 × 1.8 mm with a FOV = 360 mm, flip angle = 15°, VENC= 150 cm/s, T_R_= 4 ms, T_E_ = 2 ms. A non-invasive central blood pressure measurement was achieved approximately 30 minutes prior to each MRI session using a SphygmoCor CP system (AtCor Medical, Sydney, Australia). Systolic and diastolic pressure values were used to tune *in silico* models outlet BCs and aortic wall mechanical properties.

### 2.2. Anatomical model and finite element mesh

One pre-operative and two post-operative models, with Dacron and EW graft, were deployed in the digital twinning platform CRIMSON [25]. The embedded CRIMSON segmentation tool was employed to generate 3D geometric models of the aorta, including the supra-aortic branches, before (see Figure 1) and after grafting surgery. Each anatomical model was then discretized into tetrahedral elements, with an elements size chosen according to the results of a mesh sensitivity analysis, using CRIMSON embedded TetGen algorithm [26]. Mesh quality was evaluated assessing the *radius ratio, edge ratio* and *aspect ratio*.

**Figure 1:**
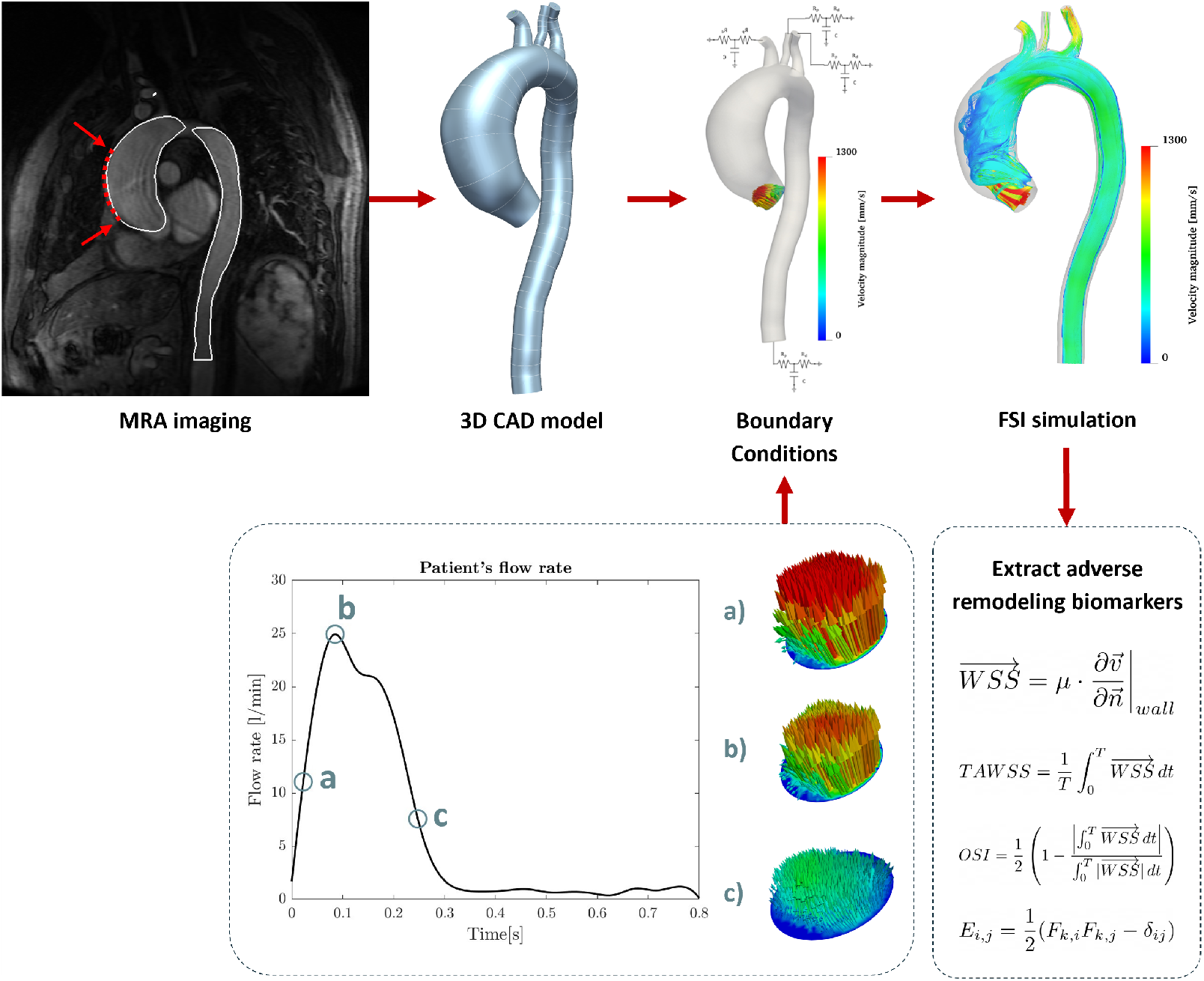
Overall framework adopted for the FSI analysis, showing the main steps: *i*) Segmentation of MRA imaging; *ii*) 3D CAD model reconstruction, *iii*) Assigning boundary conditions. Time-varying velocity field extracted from 4D flow was mapped onto the inlet plane, Windkessel model were coupled at the outlets; *iv*) Running FSI simulation.

### 2.3. Boundary conditions

#### Inlet BCs

An in-house Python code [27], was used to define PC-MRI based velocity inlet BC, discretizing the velocity field in space and time. Briefly, 4D flow data were coregistered to the reconstructed 3D geometries. Then, a radial basis function interpolation approach was used to extract velocity profiles from 4D flow data, at each time frame, and map them onto the nodes of the model target inlet surface [28]. To ensure more stable solutions, boundary velocity and backflow were set to zero. Finally, the velocity profile was up-sampled using the same time-discretization adopted for the simulations flow rate was evaluated (Figure 1, Boundary Conditions). The resulting velocity dataset was converted into a CRIMSON-readable file, to set the inflow BC.

#### Outlet BCs

A three-elements Windkessel model (3WKM), consisting of a proximal resistance R_p_, directly coupled with the 3D fluid domain, a compliance C and a distal resistance R_d_, was set as outlet BC, to account for the impact of distal vasculature [10]. The model is described by a set of ordinary differential equations (ODEs), under the assumption of uniform distribution of system variables (i.e., pressure and flow) at any time instant within any constitutive element. The characteristic coefficients (R_p_, C, R_d_) of each 3WKM were computed solving an optimisation problem related to two reduced order model (ROM) (see Figure 2), representing the patient’s aorta before and after the surgery, respectively. These ROM models provided a physics description of the aortic district under two main assumptions: *i*) The spatial variation of the pressure and flow was neglected; *ii*) The three supra-aortic branches were merged into one single branch. First, the hydraulic impedances associated with the ascending aorta (ATAA), the aortic arch (AA) and the descending aorta (DTA) were characterized with preliminary CFD simulations (see Supplementary Material). The aortic arch non-linear resistance was equally split before (R_A1_) and after (R_A2_) the only supra-aortic branch, while the linear resistances R_ATA_ and R_DA_ represent the impedance of the ascending and descending aorta section, respectively (see 2).

**Figure 2:**
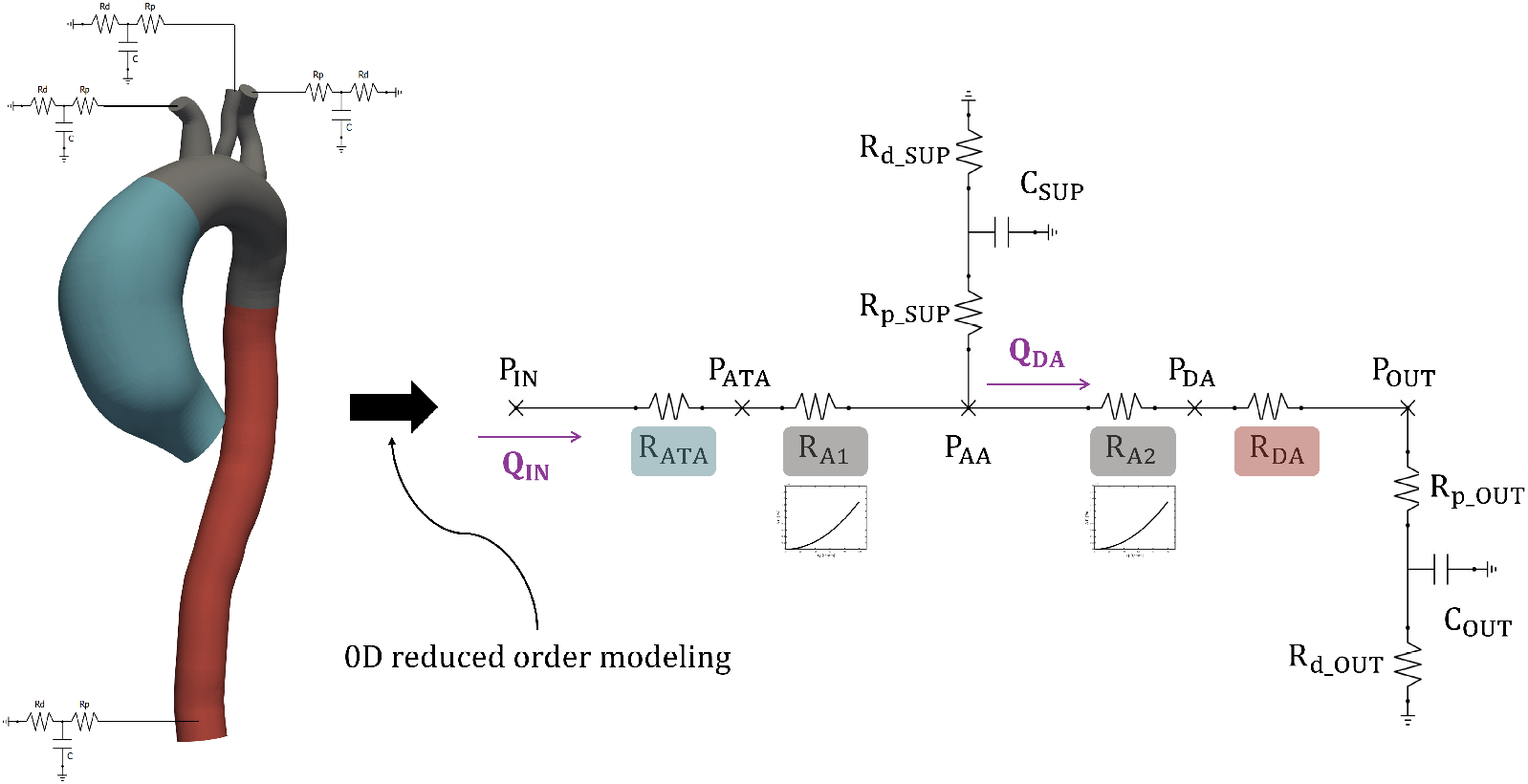
Pre-surgery configuration of the 0D lumped parameters circuit used for the tuning of outlet boundary conditions. The impedances associated with the ascending aorta, the aortic arch and the DTA were modeled with non-linear resistances, namely: *R*_*ATA*_, *R*_*A*1_-*R*_*A*2_, *R*_*DA*_.

The 3WKM coefficients of the supra-aortic branch and the DTA outlet were identified minimizing the relative squared error between:

- The patient’s analytically estimated (from the 0D ROM) inlet pressure (P_IN,*an*_) and *in vivo* measured values of diastolic (P_d_), systolic (P_s_) and mean (P_m_) blood pressure;
- The patient’s analytically estimated (from the 0D ROM) flow in DTA (Q_DA,*an*_) and the 4D flow derived corresponding quantity (Q_DA_).

By leveraging 4D flow derived inflow (Q_IN_) and combining model ODEs, inlet pressure (P_IN,*an*_) and DTA outflow (Q_DA,*an*_) can be expressed as a function of the 3WKM coefficients (see Supplementary Material).

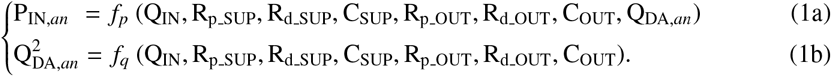

Optimization was interatively achieved in two substeps: first, Microsoft Excel (Microsoft Corporation, Redmond, WA, USA) solver was used to optimize (using the Generalized Reduced Gradient method) the RCR parameters, minimizing the DTA flow rate error; then, P_d_, P_s_, P_m_ errors were minimized. This procedure was iterated until an error below 1%, for both steps, was yielded.

Finally, the resulting RCR values for the supra-aortic branch were split between the brachiocephalic artery (BCA), the left common carotid artery (LCCA) and the left subclavian artery (LSA) based on Murray’s law. This framework was identically implemented for both the pre- and post-surgery configurations and the resulting RCR values, for both scenarios, are presented in Table 1.

**Table 1:** Parameters for the three-element Windkessel model, for both the pre- and post-surgery configurations. The units of measure are 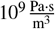 for resistance (R), and 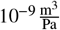 for compliance (C). *R*_*p*_=Proximal resistance, *R*_*d*_=Distal resistance.

|  | *Pre-op* |  |  | *Post-op* |  |  |
| --- | --- | --- | --- | --- | --- | --- |
| | $R_p$ | C | $R_d$ | $R_p$ | C | $R_d$ |
| BCA | 0.0773 | 3.1335 | 0.6311 | 0.0485 | 2.7698 | 0.6723 |
| LCCA | 0.1770 | 1.8042 | 1.4445 | 0.0922 | 1.8051 | 1.2780 |
| LSA | 0.0801 | 3.0622 | 0.6533 | 0.0353 | 3.4251 | 0.4890 |
| DTA | 0.0169 | 15.000 | 0.1998 | 0.0117 | 11.6556 | 0.1609 |

### 2.4. Material properties

In this section the constituive models used to capture the mechanical behaviour of blood, aortic wall and both Dacron and EW graft, are described.

#### 2.4.1. Blood

Blood was modeled as a Newtonian fluid with constant dynamic viscosity equal to μ = 4 cP, density equal to ρ = 1060 kg/m^3^ and the flow was assumed to be incompressible and laminar (during systole, Reynolds number was above 2500 only at the peak).

#### 2.4.2. Aortic wall

The aortic wall was modeled as an incompressible, elastic membrane characterized by a 5 × 5 stiffness matrix, which elements varied along the vessel centerline to capture the different response of three primary regions of interest: the ATAA, the arch and the DTA. The elements of the stiffness matrix were determined implementing the *four-fibers family* constitutive model and exploiting the theory of *small deformations superimposed on large* (SOL) [29] to linearize the material behaviour around patient’s mean arterial pressure [30]. The strain energy function reads as follow:

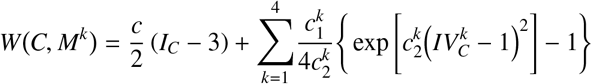

where *c*, 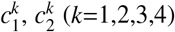 are material parameters, *C* = *F*^*T*^ *F* is the right Cauchy-Green tensor *F*, is the deformation gradient tensor, 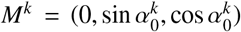 represents the orientation of the fiber families (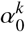 is referred to the axial direction) and *I*_*C*_, *IV*_*C*_ are the first and fourth invariant of deformation. The 8 model characteristic coefficients (i.e., *c*, 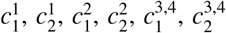 and 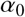) were estimated using the Levenberg-Marquardt nonlinear regression to minimize the difference between predicted and literature stress-stretch data [31, 32, 33], in absence of *ex-vivo* experimental data. As reported by Hopper *et al*. [34], linearization should occur in a ball centered in λ_*m*_, corresponding to mean pressure (*P*_*m*_). However, such stretch value was not available and to identify an equivalent stress-stretch configuration (corresponding to *P*_*m*_) we procedeed as illustrated in Figure 3.

**Figure 3:**
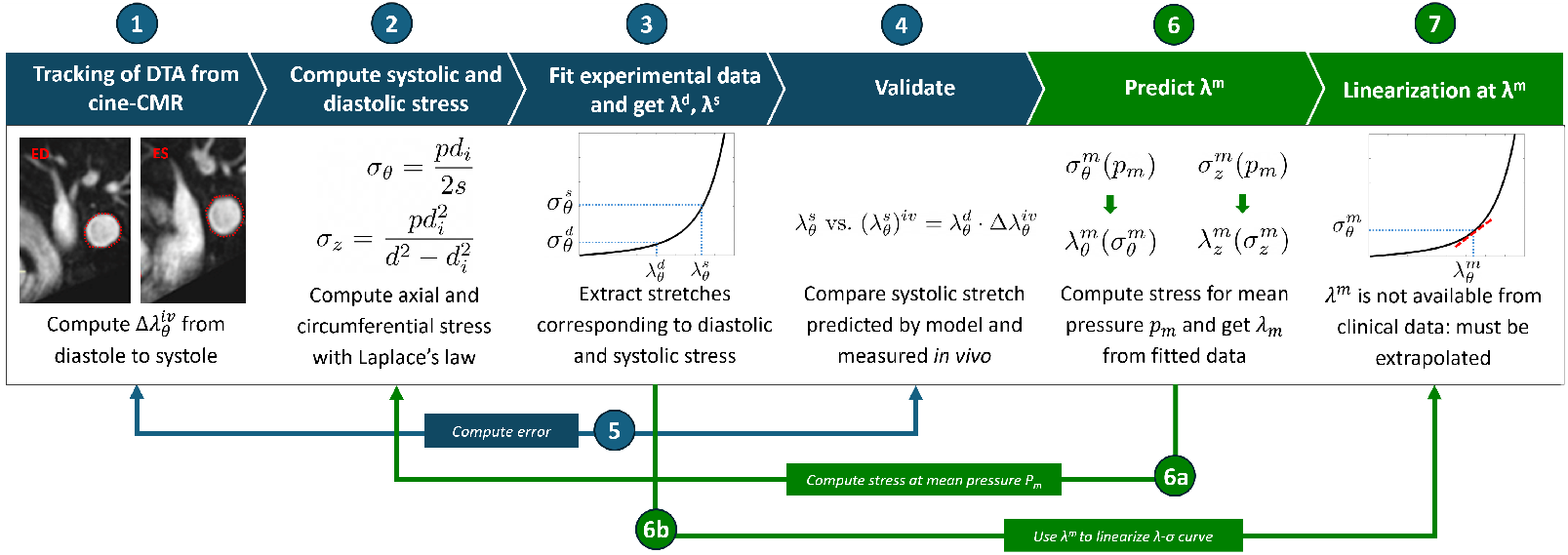
Stepwise process adopted to linearize the hyperelastic mechanical behavior of materials at average stress-stretch state.

1. The *in vivo* diastolic-to-systolic circumferential strain increment 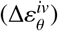 throughout the cardiac cycle was quantified from cine CMR acquisitions. The corresponding stretch value, shown in Table 2, was evaluated as:

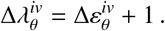
2. Circumferential and axial stresses, at diastole 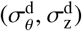 and systole 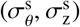, were computed from the measured P_d_ and P_s_ using Laplace’s law:

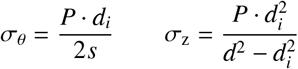

where *d*=30 mm is the vessel external diameter, *d*_*i*_ = *d* − 2*s* is the luminal diameter and the thickness *s* was assumed to be 15% of *d* [34].
3. The coefficient of the 8 parameters model reported in [31] were used to define *W* and best-fit the analytical curves representing the equibiaxial mechanical response of the arterial wall (Figure **??**). Therefore, given 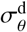 and 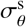 we extrapolated from the corresponding stretch value 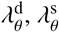 (see Table 2).
4. The stretch values obtained at the step 1 and 3 were combined to compute patient’s *in vivo* systolic circumferential stretch 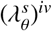, reported in Table 2 and estimated as:

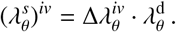
5. To ensure the robustness of framework, the error *e* between the systolic DTA diameter measured *in vivo* and estimated through 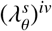 was computed:

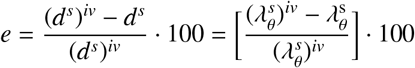

**Table 2:** Table reports, for each aortic segment in the pre-op and post-op configuration, the circumferential stretch increment measured *in-vivo* from diastole to systole, 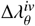 the diastolic and systolic stretch computed with the adopted consitutive model, 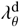 and 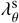 ; and the *in-vivo* estimated systolic stretch, 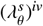. In bold are highlighted the values used for validating the pipeline described in 2.4.2. ATAA=Ascending Thoracic Aortic Aneurysm; Arch=Aortic Arch; DTA=Descending Thoracic Aorta; Dacron=Dacron graft

|  | *Pre-op* |  |  |  |  | *Post-op* |  |  |  |
| --- | --- | --- | --- | --- | --- | --- | --- | --- | --- |
| | $\Delta\lambda_\theta^{iv}$ | $\lambda_\theta^d$ | $\lambda_\theta^s$ | $(\lambda_\theta^s)^{iv}$ | | $\Delta\lambda_\theta^{iv}$ | $\lambda_\theta^d$ | $\lambda_\theta^s$ | $(\lambda_\theta^s)^{iv}$ |
| ATAA | 1.023 | 1.170 | 1.231 | 1.197 | Dacron | 1.004 | 1.319 | 1.364 | 1.324 |
| Arch | 1.065 | 1.170 | 1.231 | 1.246 | Arch | 1.086 | 1.186 | 1.237 | 1.288 |
| DTA | 1.055 | 1.179 | **1.218** | **1.244** | DTA | 1.103 | 1.190 | **1.219** | **1.314** |

An error of *e* = 2.09% for the pre-operative configuration and *e* = 7.26% for the post-operative configuration was obtained, indicating that the described pipeline is reliable. Therefore, the *four-fibers family* constitutive model proved accurate in replicating the patient’s vascular tissue response, and was consequently used to determine the mean axial stretch 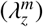 and circumferential stretch 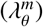 using *P*_*m*_ in Step 2. Table 2 summarizes the *in vivo* measured circumferential stretches, the analytically computed 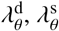, and the *in vivo* estimated 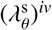.

Aortic wall stretch-stress curve was linearized around 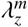 and 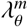, for axial and circumferential direction. Finally, the linearized anisotropic stiffness matrix ℂ_*ijkl*_ was obtained as follows, for each arotic segment:

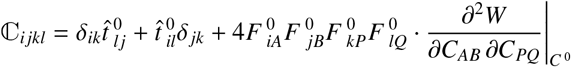

where 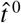, *F*^0^ and *C*^0^ are, respectively, the deformation-dependent Cauchy stresses, the deformation gradient from an undeformed state and the right Cauchy-Green tensor (see Supplementary Material).

#### 2.4.3. Dacron graft

To model the Dacron-reconstructed aortic segments, the *four-fiber family* model was implemented. Using SOL theory, the vessel wall behavior was linearized around the patient’s post-surgery mean arterial pressure configuration. Results from Dacron equibiaxial tests were retrieved from the work of Tremblay *et al*. [35], and the best-fitting parameters for the constitutive model were obtained using the Levenberg-Marquardt algorithm. The Dacron response was linearized around the blood mean arterial pressure configuration (Figure **??**). The corresponding axial 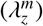 and circumferential 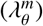 stretches were computed following the procedure described in 2.4.2, ultimately yielding the anisotropic linearized stiffness coefficients ℂ_*ijkl*_. The graft diameter and thickness were assumed to be *d* = 30 mm and *s* = 0.6 mm, respectively [35].

#### 2.4.4. Electrowritten graft

Figure 4 illustrates the workflow adopted for the EW graft characterization. The mechanical properties of the innovative device were extrapolated from uniaxial tensile tests data of PCL fibrous scaffolds reported by D’Amato *et al*. [6]. Considering the device structure, the *two-fibers family* constitutive law was adopted, which assumes identical and symmetrically oriented (with respect to device axis) fibers. Therefore, the strain energy function reads as:

**Figure 4:**
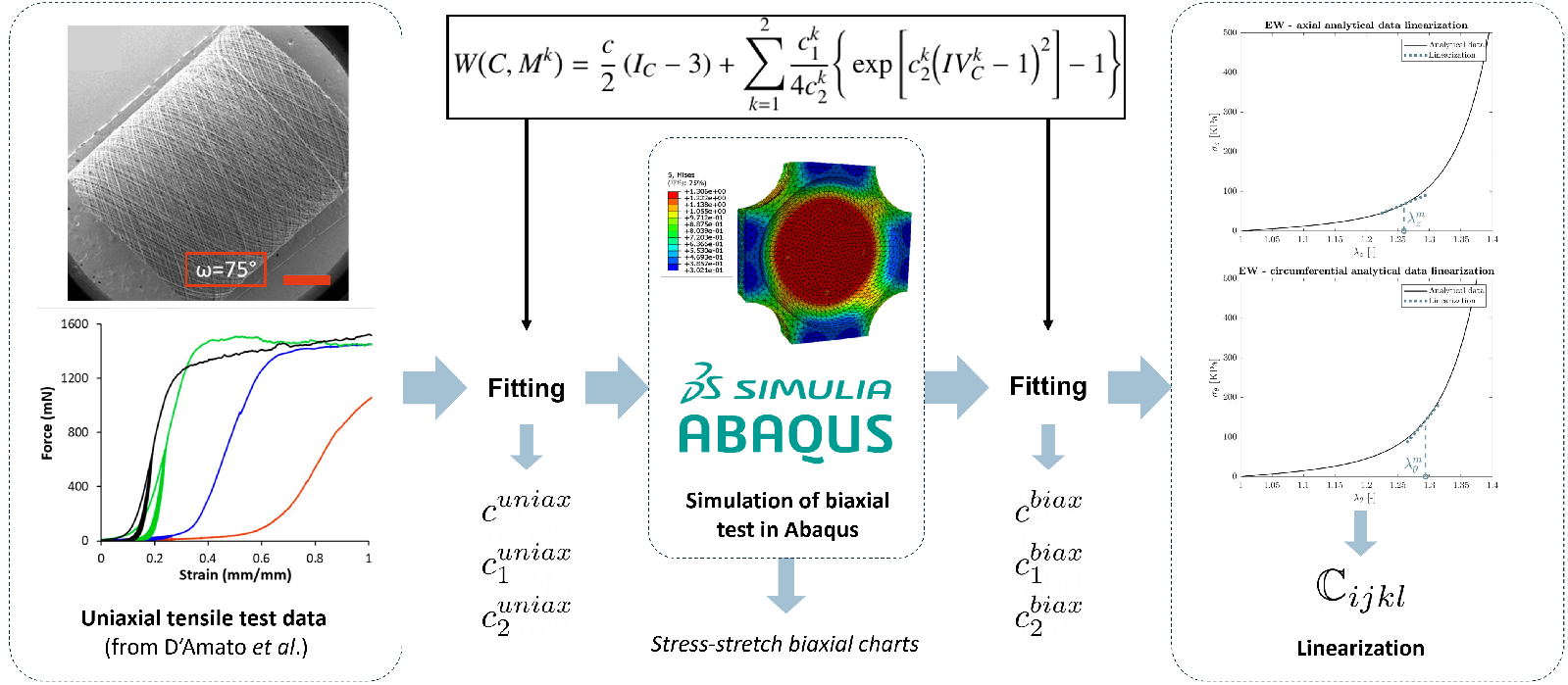
Workflow followed for the virtual EW graft mechanical behaviour characterization.

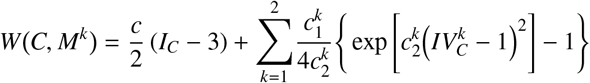

The coefficients of the model (i.e., *c, c*_1_, *c*_2_) were estimated best-fitting the uniaxial tests data for a scaffold with winding angle ω = 75°, which mimicks large vessel structural characteristics, under the assumption of incompressible behavior and ideal uniaxial tensile state (i.e., σ_*r*_ = σ_θ_ = 0).

Since the available experimental data were inadequate to represent the anisotropic material response, an equibiaxial tensile test up to 30% of nominal strain was simulated in Abaqus/Standard (Simulia, Dassault Systèmes, Providence, RI, USA) for an incompressible square patch (details of the simulation are available in the Supplementary Material). Its mechanical response was modeled with the anisotropic hyperelastic Holzapfel-Gasser-Ogden constitutive law, fed with the previously best-fitted parameters (i.e. *c, c*_1_, *c*_2_). Stretch-stress curves were extracted from the simulation output and used to best-fit again the *two-fibers family* parameters using Lavemberg-Marquardt minimization method. The resulting curve was linearized around post-operative mean pressure (*P*_*m*_) (see Figure **??**). Mean pressure axial 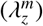 and circumferential 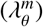 stretches were estimated using the same approach described in 2.4.2, assuming the following geometrical features: *d*= 30 mm and *s*= 1.4 mm [6]. The alternative post-operative scenario was modeled substituting the traditional Dacron graft with the virtual EW device. Finally, linearized anisotropic stiffness coefficients ℂ_*ijkl*_ of the EW graft were computed and fed to CRIMSON FSI code. Table 3 sums up the resulting axial (ℂ_*zzzz*_) and circumferential (ℂ_θθθθ_) components of the stiffness matrix for the baseline ATAA, the Dacron graft and EW graft.

**Table 3:** Linearized anisotropic stiffness coefficients in the axial (ℂ_*zzzz*_) and circumferential (ℂ_θθθθ_) direction for the pre operative ascending thoracic aorta, Dacron and EW graft

| | $\mathbb{C}_{zzzz}$ [MPa] | $\mathbb{C}_{\theta\theta\theta\theta}$ [MPa] |
| --- | --- | --- |
| **ATAA** | 0.335 | 0.570 |
| **Dacron** | 1.791 | 2.248 |
| **EW** | 0.648 | 1.310 |

### 2.5. Model Validation

Model validation was achieved qualitatively and quantitatively comparing simulations results and 4D flow data on six planes identified along aorta centerline: P_1_, P_2_, P_3_ in the ascending aorta and P_4_, P_5_, P_6_ in the descending section, as depicted in Figure **??**.

Velocity magnitude contours and streamlines were qualitatively compared: an in-house Python algorithm [27] was used to project 4D flow velocity data onto each plane of interest, whereas 4D flow streamlines were extracted in Paraview. For quantitative validation, ascending and descending aorta flow rates, evaluated through 4D PC-MRI and FSI models, were compared. In addition, the aortic pressure, measured before MR scanning, was compared to the inlet pressures predicted by the FSI analysis.

### 2.6. Biomarkers of adverse remodeling

The risk of adverse remodeling induced by the grafting procedure was assessed using three hemodynamic biomarkers: the wall shear stress (WSS), the time-averaged WSS (TAWSS), and the oscillatory shear index (OSI). Altered values of these indeces can trigger different pathways that lead to atherosclerosis initiation. High value of WSS, and low WSS associated with high OSI, can both induce atherogenesis [36]. WSS is defined by Newton’s law:

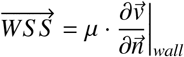

Where 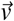 is the velocity, 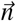 is the normal direction to the wall, μ is the dynamic viscosity.

The TAWSS assesses the average shear stress experienced by the vessel over a cycle of period *T*. It is defined as:

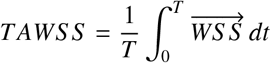

OSI measures the influence of the oscillatory component of WSS. Time variations of WSS magnitude and direction have been shown to have a significant impact on vessel health, playing a crucial role in triggering the adverse remodeling process [37]. Atherogenic process is triggered when WSS and OSI approach 0 and 0.5, respectively [36]. OSI is defined as:

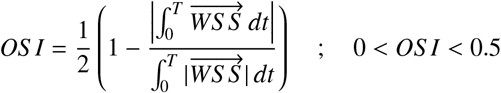

Additionally, as a structural biomarker, the maximum principal strain (i.e., the first eigen-value of the Green strain tensor) was analyzed. Since the CRIMSON FSI code outputs only the displacement field, an in-house Python code was used to extract the resulting strain field [38].

First, the distributions of these biomarkers were assessed over the whole aorta for all computational models and compared to each other. Then, the analysis focused on the proximal descending aorta (Figure 10a), where graft-induced adverse remodeling processes could initiate.

### 2.7. Statistical analysis

Statistical analysis was conducted using GraphPad Software (San Diego, California, USA) to compare the distribution of each field in the DTA between baseline and post-op scenarios. To enable a per-point comparison of field distributions on different baseline and post-op meshes, a nearest-neighbor-based interpolation of the post-op geometry was performed after coregistering the pre-op models. A paired *t-test* was used for per-point analysis if the data passed the normality test; otherwise, a paired Wilcoxon test was applied. Statistical analysis was performed for the following result combinations: *(i)* Pre vs. Post-Dacron; *(ii)* Pre vs. Post-EW; and *(iii)* Post-Dacron vs. Post-EW. Finally, the statistical differences among the three scenarios were evaluated using a paired one-way analysis of variance (ANOVA) test.

## 3. Results

Two cardiac cycles were simulated on a 12-cores Processor AMD Ryzen 9 5900X 3.70 GHz machine with 64 GB of RAM. A complete simulation required ≈ 3 to 4 days. The results from the last cardiac cycle are described in this section.

### 3.1. Model validation

#### Qualitative Comparison

The results of the FSI simulations were qualitatively compared to 4D flow data using streamlines and velocity magnitude plots across six cross-sections along the ATAA and DTA (see Figure 6). Figure 5 shows the systolic streamlines extracted from both 4D flow and FSI simulations. In the baseline model, due to the presence of the aneurysm, vortical streamlines were observed at the intrados of the ATAA. After surgery, the flow in the ATAA tract resulted straightened.

**Figure 5:**
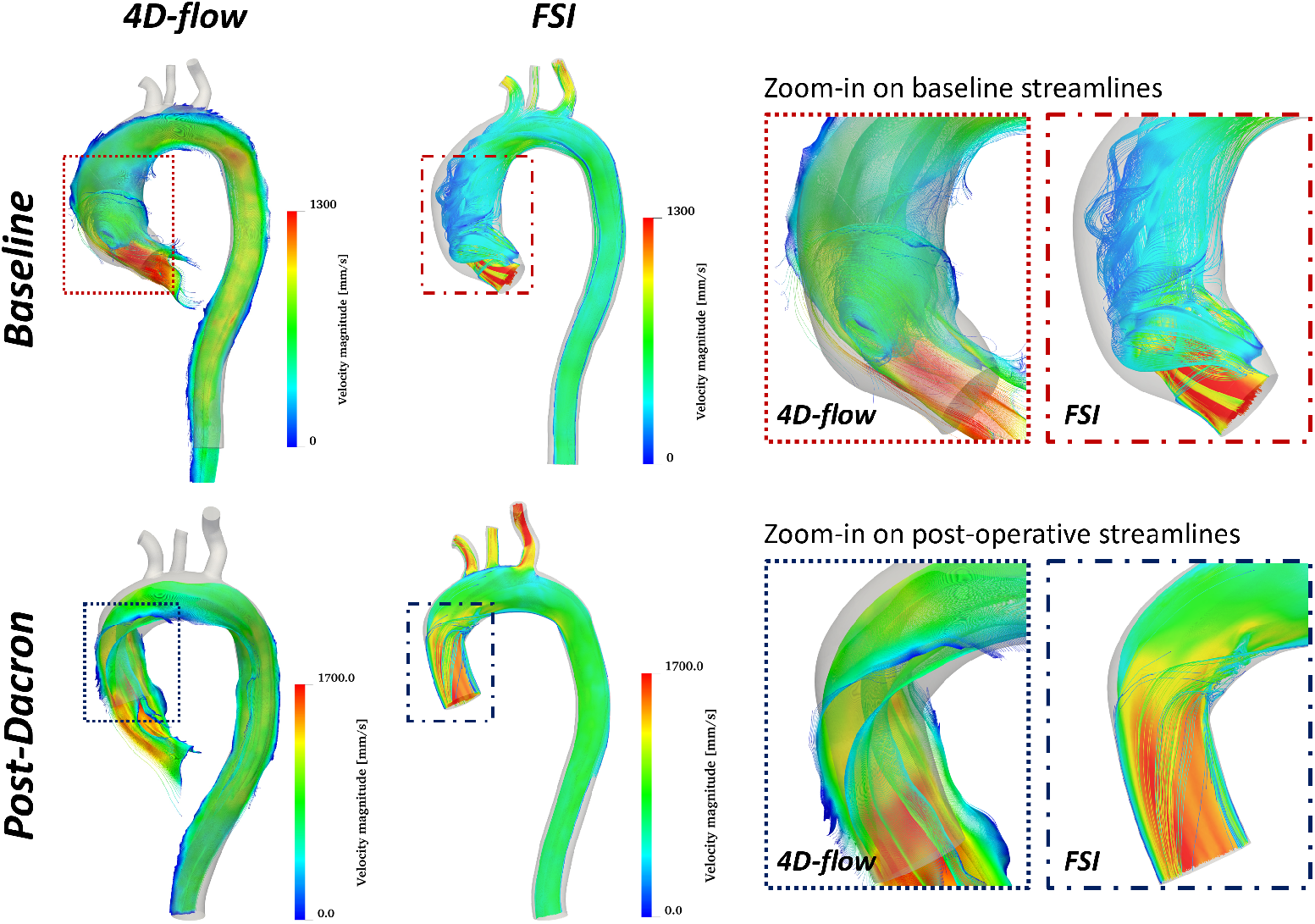
Comparison of streamlines extracted from 4D-flow sequence and FSI simulations for both the baseline and actual post-operative model. A zoom-in on the ascending aorta region is showed on the right for each model.

**Figure 6:**
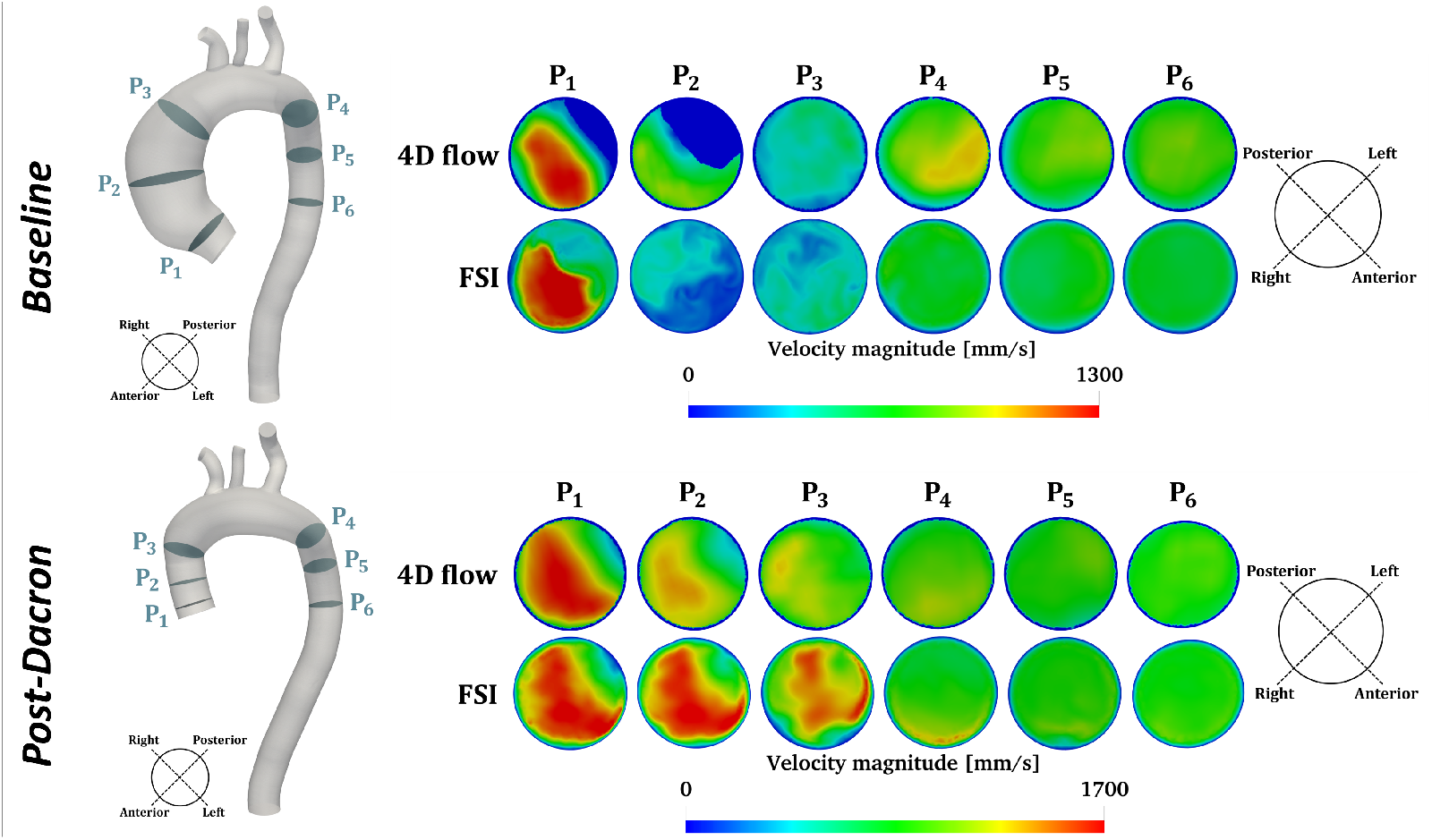
Qualitative comparison of velocity contour, on six cross sections along the centerline of the aorta, extracted from 4D-flow sequence and FSI simulations for both the baseline and actual post-operative model.

Figure 6 depicts the velocity magnitude field over six cross section along the aorta. The DHT was generally able to predict the velocity pattern obtained from 4D flow. In the baseline model, on P_1_, the velocity profile peak was located in the same region both in 4D flow and in the FSI result. On P_2_ the two contours exhibited the largest visual mismatch in terms of distribution and magnitude. On P_3 *to* 6_ a good agreement can be observed between 4D flow and DHT in terms of both velocity profile pattern and magnitude. In the post-operative configuration, the comparison between 4D flow and FSI yielded a worse agreement in the ATAA, with respect to the previous scenario. In the DTA cross sections, a good agreement was observed between 4D flow and the DHT in terms of velocity pattern.

#### Quantitative Comparison

The outflow rate in DTA was compared between 4D flow and the DHT model. In the baseline case (Figure 7a), the DHT accurately reproduced the flow rate waveform (mean absolute error, *MAE*=1.39 L/min), yielding a Spearman correlation coefficient *r*= 0.6050, p<0.0001. The largest difference occurred at the systolic peak (relative squared error, RSE=5.28%). In the post-operative configuration (Figure 7b) a *MAE*=0.99 L/min, with *r*= 0.8693, p<0.0001 was obtained. In this scenario, the systolic peak flowrate in the DTA was precisely predicted (RSE=0.33%).

**Figure 7:**
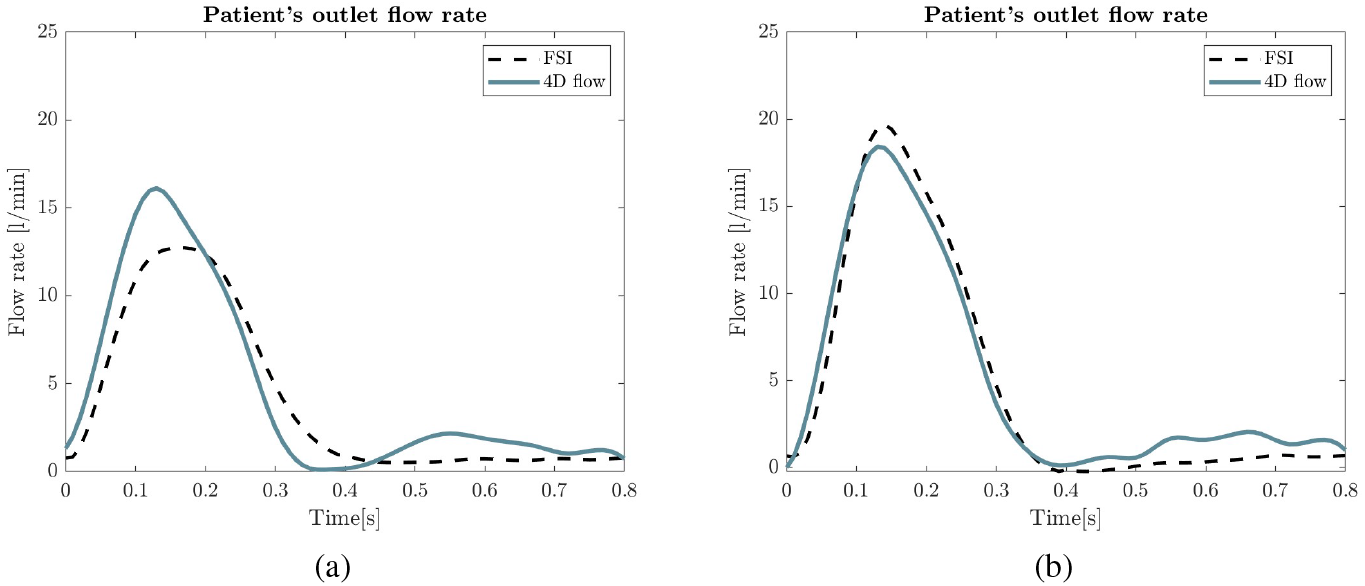
Comparison of outflow in the descending aorta extracted from 4D flow and FSI simulation, in the baseline (a) and post-operative (b) configuration.

Finally, clinical acquisitions of central blood pressure were compared versus the inlet pressure predicted by the FSI model. Before surgery, a systolic (SP) and diastolic (DP) pressure were 132 mmHg and 85 mmHg, respectively. A SP= 120 mmHg and DP=75 mmHg were computed from the FSI model at the aortic inlet. Thus, a physiological pressure range was reproduced, slightly underestimating patient’s pressure (by ≃ 9% and ≃ 12%). After surgery, measured SP and DP were 137 mmHg and 96 mmHg. The values predicted by the FSI model at the aortic root were SP=125 mmHg and DP=88 mmHg. Similarly to baseline model, a slightly lower pressure range was reproduced (by ≃ 9% and ≃ 8%).

### 3.2. Comparison of baseline v. post-surgery scenarios

The comparison between the pre and post surgery configuration was based on the distribution and peak values analysis of the biomarkers described in Section 2.6. The analysis also included the alternative post operative scenario, in order to investigate the capability of more compliant EW graft to mitigate hemodynamics anomalies due to traditional surgery. Indeed, those indices are considered extremely useful tools in the clinical follow up, since they suggest whether and where the graft-induced adverse modeling process may origin [7, 36]. All values are reported as *median* [*10*^*th*^ *to 90*^*th*^ *interquartile range*].

#### Maximum WSS

The WSS field on the whole aorta and, at the systolic peak, is illustrated in Figure 8, for the baseline model and for the post-surgery with Dacron and EW graft. The pre-op scenario was characterized by an intensification of WSS magnitude in the ATAA, immediately after the aortic inlet, due to the highly skew inlet velocity profile. In the baseline model, excluding the supraortic and BCs regions, the WSS was on average 3.82 Pa [1.49; 6.16]. In both post-op scenarios, an overall increment of WSS was observed (the average WSS was 8.62 Pa [3.10; 22.95] for DG and 8.14 Pa [3.13; 22.50] for the EW graft model), with peak values that interested mainly the ascending aorta and the intrados of the proximal descending aorta.

**Figure 8:**
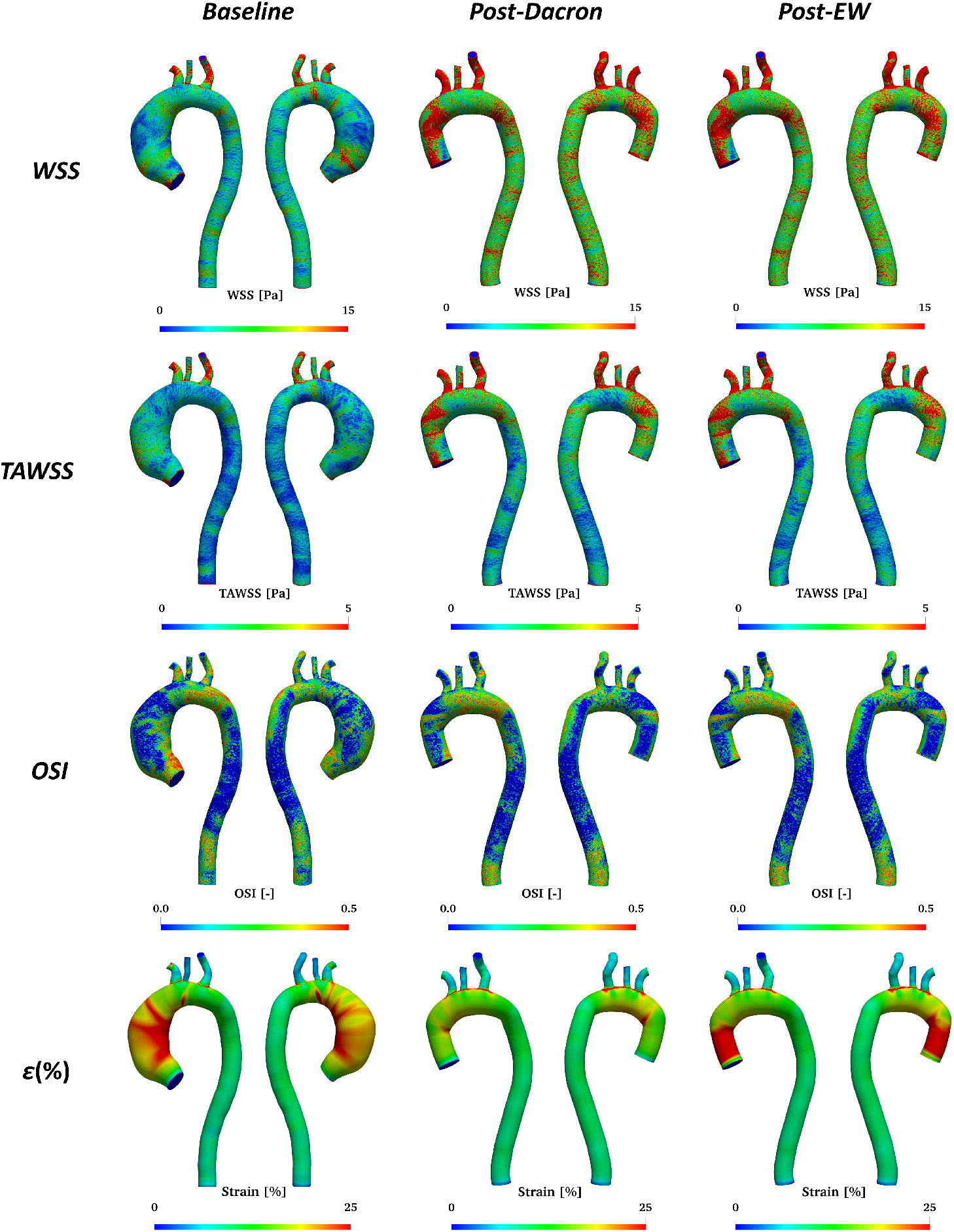
From top to bottom row: resulting WSS, TAWSS, OSI and maximum principal strain fields of the FSI analysis. For each field is reported, from left to right, the baseline, the actual post-surgery with Dacron graft and the innovative post-surgery with electrowritten graft.

#### TAWSS

The TAWSS field (Figure 8) showed a similar pattern to systolic WSS. From baseline to post-operative model, a visible increase in TAWSS along the aorta is visible: the average value in the aorta was 0.89 Pa [0.30; 2.48] at baseline and, 2.28 [0.45; 4.86] Pa and 1.45 Pa [0.44; 4.78] in the post-operative model with Dacron and EW graft, respectively. After surgery, the average stress exceeded the physiological value of 4-5 Pa in many region, identifying arae where adverse remodeling could initiate.

#### OSI

The third row in Figure 8 shows the OSI field for the three models. In the baseline model, OSI reached high value (i.e., ≈ 0.5, indicating an increased atherogenic process starting risk [36, 37]) in the ATAA, close to the sinotubular junction, where vortical structures are visible in the streamlines, and in the arch. In both post-surgery scenarios several region at high OSI value were found, mostly in the arch an proximal DTA. On average, OSI resulted 0.17 [0.01; 0.43], 0.16 [0.01; 0.42], 0.16 [0.01; 0.43] for the baseline, Dacron and EW graft models, respectively.

#### Maximum principal strain

The maximum principal strain field at the systolic peak is depicted in the last row of Figure 8. Before surgery strain was on average 3.10% [0.84%; 6.77%], maximum values occurred along the ascending aneurysmatic aorta, at the intrados. In both grafted models, the highest deformations were observed at the anastomosis and along the aortic arch. On average, strain resulted 1.96% [0.84%; 3.82%] and 2.13% [0.85%; 4.02%] for Dacron and EW graft, respectively. In the distal aorta, an increase in strain magnitude compared to the baseline is visible in both post-operative scenarios. As expected, the more compliant EW device experienced a larger deformation compared to the Dacron graft, resulting in a slightly higher strain range, closer to baseline model.

#### 3.2.1. Focus on the proximal descending aorta

Hemodynamics changes, associated to ATAA graft implantation, mainly affects the descending aorta [3]. Thus, a deeper analysis was performed focusing on the proximal DTA. In Figure 9 the per-point difference (i.e., Δ *f* (**x**) = *f*_*post*_(**x**) − *f*_*pre*_(**x**)) of maximum WSS and strain fields, between the actual post-op scenario and baseline. Visually, a qualitative increment of both biomarkers along the proximal descending aorta after surgery, focused at the intrados, was observed. On average, WSS and strain increment was +8.15 Pa [+0.02; +30.98] and +2.61% [+1.36%, 5.00%], respectively.

**Figure 9:**
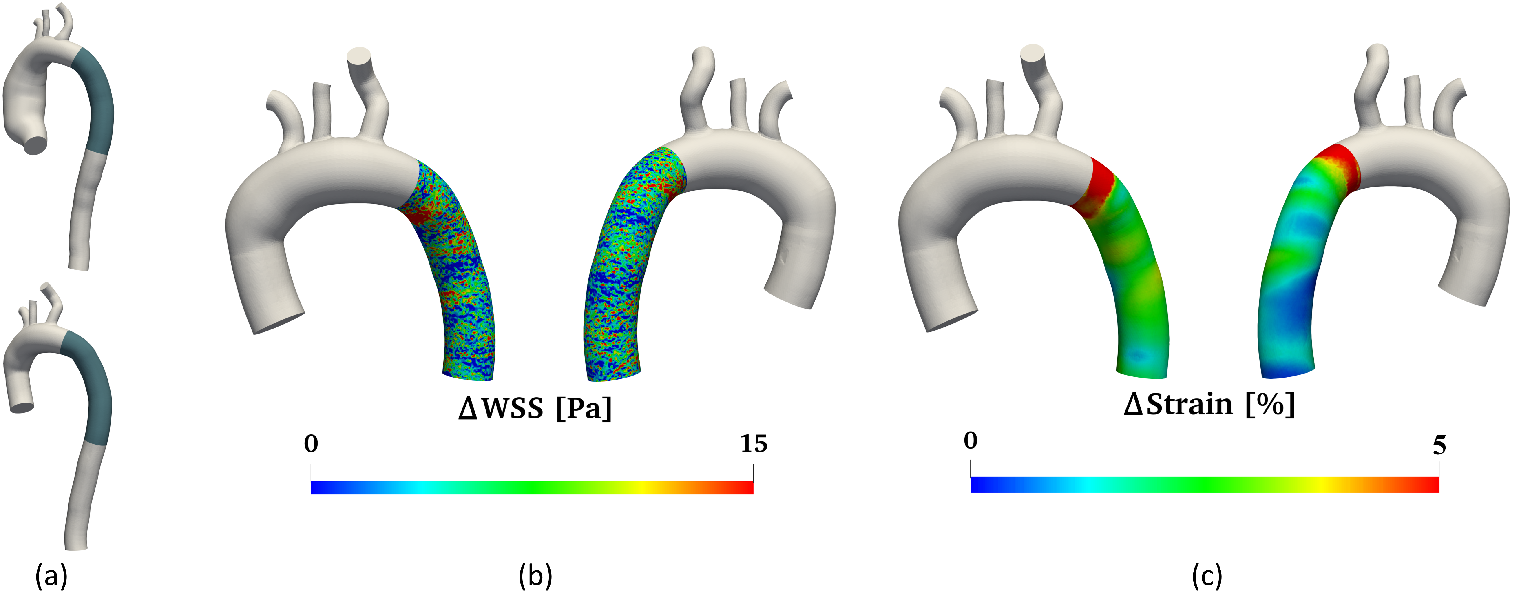
Proximal descending thoracic aorta region of analysis (a). Per-point difference of maximum WSS (b) and principal strain (c) between the actual post-operative and baseline scenario.

The same comparison was conducted between DG and EW post operative configurations Figure 10 depicts the per-point difference of the WSS fields in the DTA (i.e., Δ*WS S* (**x**) = *WS S* _*DG*_(**x**) − *WS S* _*EW*_ (**x**)). WSS resulted higher in the DG scenario, by +0.6 Pa [-0.09; +2.83], on average.

**Figure 10:**
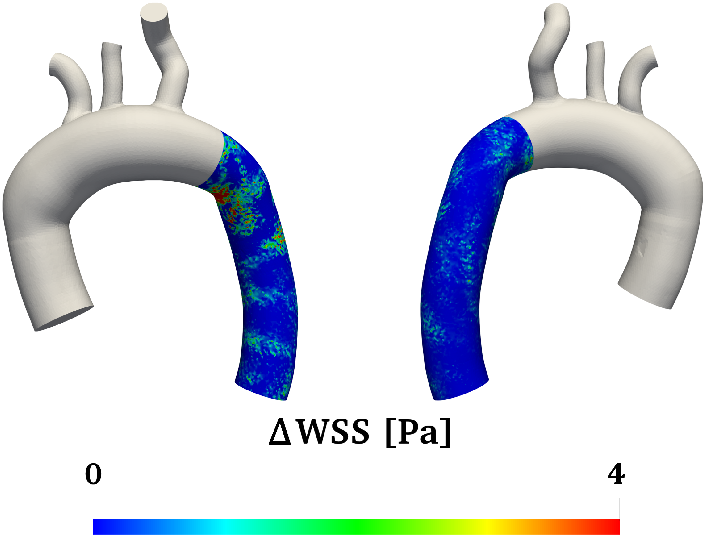
Per-point difference of maximum WSS between the actual (Dacron) and the innovative (electrowritten) post surgery scenarios.

Statistical analysis focused on WSS, TAWSS, OSI and strain fields for three simulations: *(i)* Pre vs Post-Dacron, *(ii)* Pre vs Post-EW and *(iii)* Post-Dacron vs Post-EW. Since no dataset passed the normality test, a paired Wilcoxon t-test was performed. A statistically significant difference (*p* < 0.0001) was found in the comparison of each biomarker distribution (Figure 11) for each pair of scenarios, except for the OSI comparison between the Dacron and EW cases (*p* > 0.05). Both post surgery scenarios exhibited considerably higher WSS, TAWSS, OSI and strain with respect to the baseline. Conversely, the comparison between Dacron and EW graft generally showed slightly lower mean values in the EW case. The Friedman ANOVA test between the three scenarios resulted in a marked statistically meaningful difference (*p* < 0.0001) for all the analyzed biomarkers.

**Figure 11:**
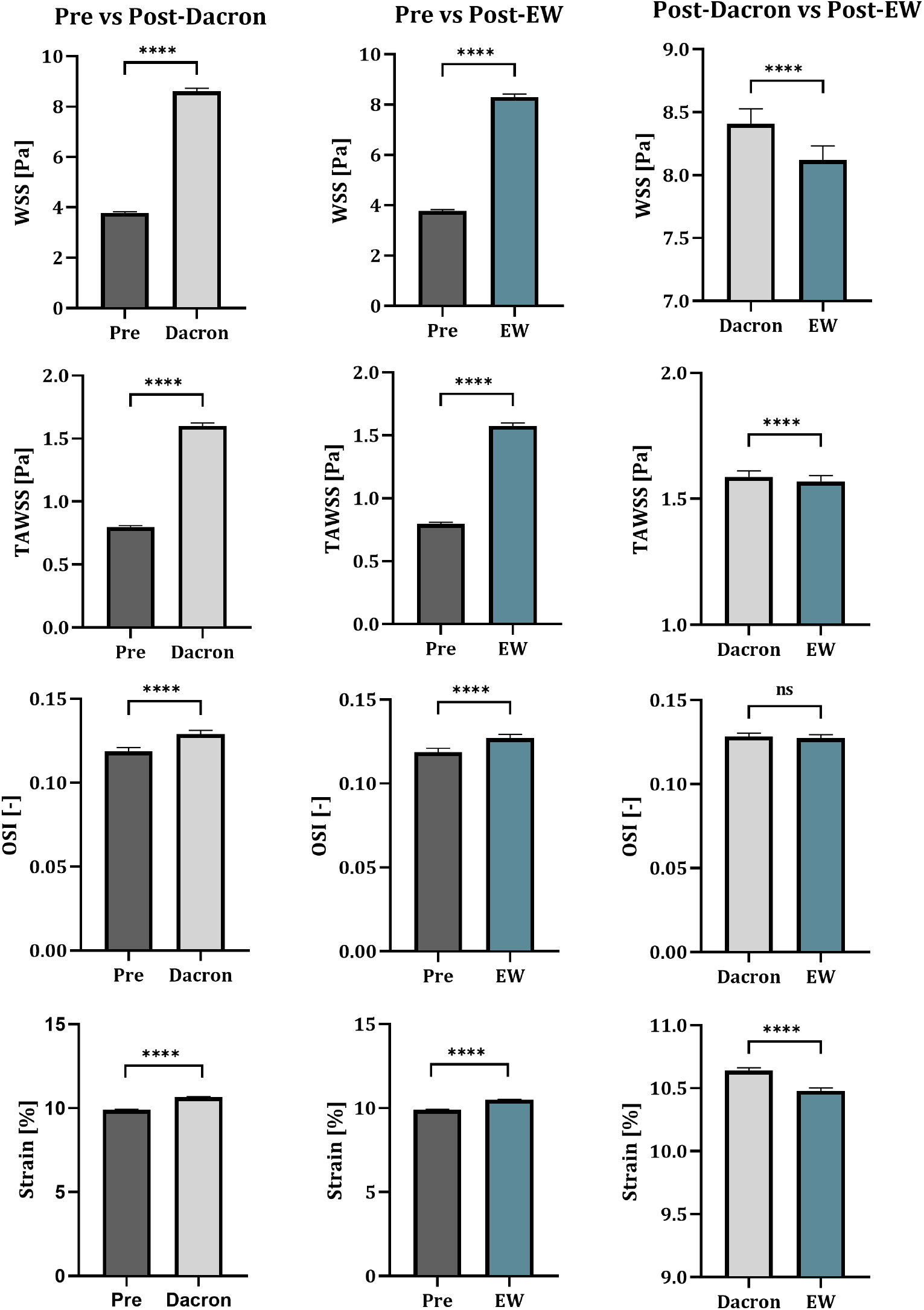
Quantitative comparison of systolic peak distributions of WSS, TAWSS, OSI and strain along the proximal descending aorta for three simulations results combinations: at the left pre vs post-Dacron, at the center pre vs post-EW and at the right post-Dacron vs post-EW. In each bar graph the mean value, its 95% confidence interval and the paired Wilcoxon t-test result are reported. *p-value* legend: ns = p > 0.05, * = p < 0.05, ** = p < 0.01, *** = p < 0.001, **** = p < 0.0001

## 4. Discussion

Aortic Dacron grafting is an effective treatment for aneurysm, but introduces a compliance mismatch between the native tissue and the prosthesis, potentially causing biomechanical alterations [4], that may lead to adverse remodeling. The predictive capabilities of physics-based or data-driven digital human twins are a topic of growing interest in the pursuit of personalized medicine. In this study, a high-fidelity virtual replica (i.e., a DHT) of a patient with ATAA was created to investigate changes in blood fluid dynamics following ascending aorta grafting. Non-linear arterial wall behavior and anisotropy were included in the constitutive model. Patient-specific inlet and outlet boundary conditions were imposed to replicate *in vivo* flow conditions. A DHT representing *i*) the pre- and *ii*) the actual post-surgery scenarios was created and validated against clinical data. Then, a third virtual post-surgery model using a different prosthesis was simulated, to predict the outcome of a different surgery strategy for mitigating the adverse effect of the Dacron prosthesis.

### 4.1. Reliability of the DHT

To assess the DHT’s reliability, the results of numerical models were compared against *in vivo* 4D flow clinical data. Streamlines and in-plane velocity patterns were qualitatively similar in 4D flow and FSI datasets, both for the baseline and Dacron graft model (Figures 5, 6). Outflow rates were slightly underestimated in the baseline model (RSE = 5.28%, *r*=0.605) whereas a good accuracy was achieved in the post-surgery model (RSE = 0.33%, *r*=0.869). The discrepancies in the DTA may stem from the mechanical characterization of the arterial wall in the ATAA, that was based on literature data. However, pathological tissues can exhibit very heterogeneous mechanical properties, difficult to capture without *in-vitro* testing of the specific tissue or more sophisticated imaging analysis. Hopper *et al*. [34] remark the impact of arterial stiffness on pulse wave velocity propagation and hemodynamics within the vessel, stressing that proper tissue characterization is crucial for high fidelity *in silico* models. Overall, the comparison between FSI results and 4D flow data, confirms the ability of the DHT to replicate patient’s biomechanics.

### 4.2. Comparison pre vs post surgery

Hemodynamics anomalies induced by the endovascular grafts were assessed based on the WSS, TAWSS, OSI and principal strain. The qualitative comparison (Figure 8) of the WSS fields showed different patterns and magnitude values, confirming an overall increment of WSS after grafting, consistently with prior work by Gaudino *et al*. [4]. Maximum stresses were observed at the anastomosis and in the proximal descending aorta intrados (≈10 Pa). These results aligns with previous findings both in terms of peak stress location and magnitude [5]. The per-point comparison (9) between the baseline, the actual post-surgery (Dacron graft) and the innovative post-surgery (EW graft) yielded a significant increment of stresses after grafting (*p*< 0.0001), confirming the impact of the surgery on biomarkers of adverse remodeling risks. Indeed, when endothelial cells are subjected to intense mechanical stimuli, the atherogenic process is triggered [39, 40]. The systolic WSS field obtained with the Dacron and EW graft models did not show qualitative remarkable differences: the per-point comparison, illustrated in Figure 10, showed moderate variations. The statistical analysis reported in Figure 11, pointed out a ≈3.42% lower mean value (*p*< 0.0001) in the virtual device scenario, indicating that fibrous EW graft could mitigate the risks associated to the traditional grafting.

Similar considerations extend to the TAWSS analysis. The TAWSS field showed an overall magnitude increment due to prosthesis implantation (≈2.56 and 1.62 folds for Dacron and EW graft, respectively). The quantitative comparison exhibited a relevant difference between all configurations (Figure 11) and a ≈1.15% mean value reduction for the innovative surgical scenario.

In the post-operative models, the OSI values were close to 0.5 in proximal descending aorta. In their study, Ku *et al*. [36] reported that OSI close to 0.5 is critical for the atherogenic process initiation: in these terms the proximal DTA could be identified as the most susceptible area to the adverse remodeling process, as described in previous works [38, 5]. No meaningful difference (*p*> 0.05) observed between OSI fields in the Dacron graft and EW graft models. As a consequence, in both grafted cases the atherogenic triggering risk is not negligible in that area.

Principal strain peaks were located at the ATAA intrados, in the baseline configuration, and at the anastomosis and along the aortic arch in both post operative scenarios with peak values of ≈20%. These results align with previous works [16, 41, 38]. Focusing on the DTA, an overall increment of strain (+2.61%, *p*< 0.0001) can be observed after grafting, consistently with the hypothesis that the compliance mismatch between the native aorta and the graft induce an augmented distal energy delivery [4]. The increased distensibility of distal aorta after grafting is confirmed by the analysis of cine-CMR data [42]. However, even if the EW graft behaved as an elastic reservoir, deforming more than the DG does, the relative distal strain reduction was moderate (≈1.51%) with respect to the actual post-surgery scenario, despite being statistically relevant.

Consequently, despite solution EW potential as an alternative fabrication technique for prosthetic devices, it currently demonstrates limited beneficial effects on the altered downstream vascular fluid dynamics. Therefore, while this innovative technique holds promise, it requires further investigations and studies to be perfectly tuned to patient specific needs.

### 4.3. Limitations and future developments

A major limitation in the present work relates to the innovative graft mechanical characterization: since experimental biaxial tests on the EW specimen were not available, its response was simulated in Abaqus, extrapolating the constitutive model from experimental uniaxial tests. Our future joint research with the group that fabricated the EW graft [6] aims to include a more comprehensive characterization of the device in the FSI protocol, conducting biaxial testing and directly extrapolating the constitutive model from experimental results. Additionally, this work investigated a case study to provide a proof-of-concept for the feasibility and robustness of the described framework to implement a DHT for personalized planning of ATAA surgery. The study is not based on a large cohort and the number of cases must be expanded. Given the promising results of this case study, we are currently enrolling new patients treated for ATAA, at the same hospital that provided the subject for this study and simulating these cases with the same workflow. Given the computational effort required to simulate each patient, we aim to train and integrate a geometric deep learning network to infer the field of interest (e.g., stress and strain) along the cardiac cycle, for a real-time assessment of the stress state in the patient, making our DHT both physics-based and data-driven.

## 5. Conclusions

In this study, a digital high-fidelity replica of a real-world patient was developed using an FSI-based methodology to comprehensively investigate the impact of compliance mismatch, due to proximal aneurysm grafting, on DTA hemodynamics and biomechanics. The DHT was refined to account for different aortic segment wall responses by adopting population-specific properties and utilizing a strain energy function that captures the non-linear and anisotropic behavior of the material, thus representing the complexities of vascular biology. Patient-specific flow and resistance BCs, were adopted. The model was validated against clinical data and 4D flow acquisitions, achieving an overall good agreement, ensuring reliable reproduction of the patient’s vessel *in vivo* biomechanics, and enabling the prediction of biomarkers associated with the development of adverse outcomes. We used our DHT to compare the actual post-surgery scenario, involving a Dacron-reconstructed ascending aorta, with a virtual correction using a novel electrowritten prosthesis. The results showed moderate but promising beneficial effects of more compliant fibrous devices on the patient’s distal hemodynamics.

## Data Availability

All data produced in the present study are available upon reasonable request to the authors

