## Supplementary material for "Deployment of a digital twin using the coupled momentum method for fluid-structure interaction: a case study for aortic aneurysm": CBM_supplementary.pdf

*Appendix A: Tuning of the boundary conditions*

The hydraulic impedances associated with the ascending aorta (ATA, $R_{ATA}$), the aortic arch ($AA$, $R_{AA}$), and the descending aorta ($DTA$, $R_{DA}$) were characterized with preliminary steady CFD simulations. Five CFD simulations were executed with increasing inlet flow $Q_{IN}$ from $5\, l/min$ to $25\, l/min$. Pressure drops and flow rates for ATA, AA, and DTA were extracted from the simulations, and the pressure drop-flow rate curves were reconstructed. The curves of the ATA and DTA were fitted with a linear law to obtain $R_{ATA}$ and $R_{DA}$, respectively. The curve of the AA was fitted with a second-order law to obtain $R_{AA}$. $R_{AA}$ was equally split in $R_{A1}$ and $R_{A2}$.

An optimization process identified the remaining components (i.e., 3WKM coefficients for the supra-aortic vessels, $R_{p,SUP}, R_{d,SUP}, C_{SUP}$, and the outlet, $R_{p,OUT}, R_{d,OUT}, C_{OUT}$) of the equivalent circuit of the aorta (Figure 1). The six equations used for optimizing the circuit are as follows:

$$I)\ P_{IN} - P_{ATA} = Q_{IN} \cdot R_{ATA}$$

$$II)\ P_{ATA} - P_{AA} = a \cdot Q_{IN}^2 + b \cdot Q_{IN} + c$$

$$III)\ P_{AA,i+1} = \frac{\Delta t}{C_{SUP}} \left[ Q_{SUP,i} - \left( \frac{P_{AA,i} - R_{p,SUP} \cdot Q_{SUP,i}}{R_{d,SUP}} \right) \right]$$

$$IV)\ P_{AA} - P_{DA} = a \cdot Q_{DA}^2 + b \cdot Q_{DA} + c$$

$$V)\ P_{DA} - P_{OUT} = Q_{DA} \cdot R_{DA}$$

$$VI)\ P_{OUT,i+1} = \frac{\Delta t}{C_{OUT}} \left[ Q_{DA,i} - \left( \frac{P_{OUT,i} - R_{p,OUT} \cdot Q_{DA,i}}{R_{d,OUT}} \right) \right]$$

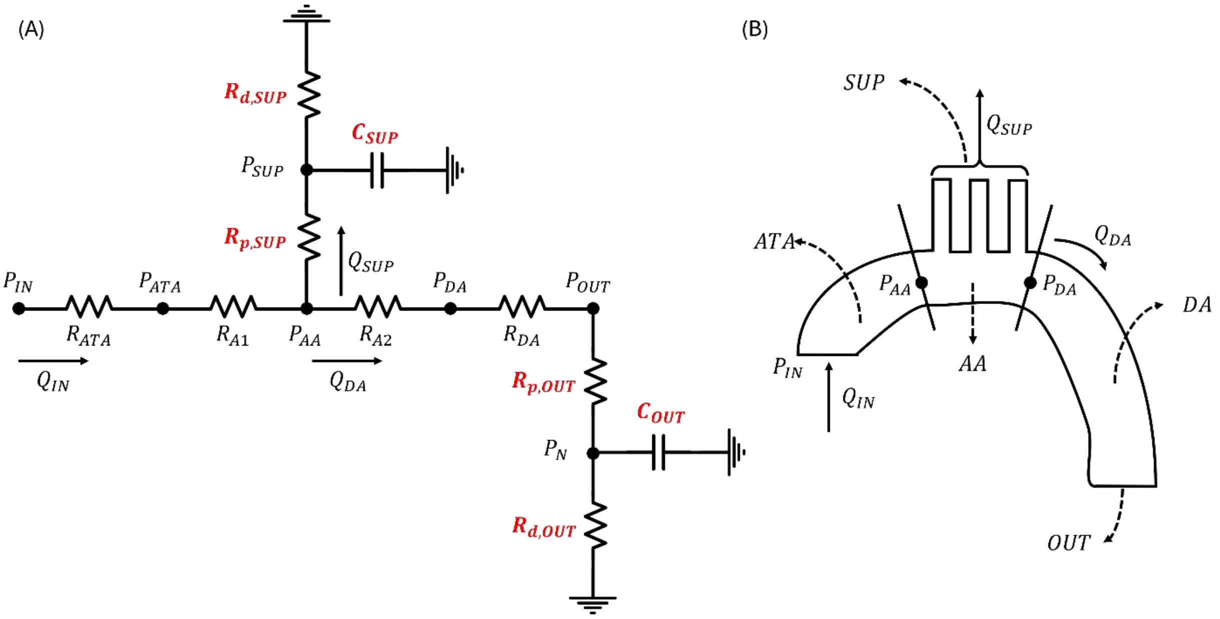


Figure 1: A) Pre-surgery configuration of the 0D lumped parameters circuit used to tune outlet boundary conditions (in red are the unknowns identified during the optimization process.). B) Schematical representation of the pre-surgery configuration of the aorta.

*Appendix B: Anisotropic stiffness matrix of the aortic wall*

The linearized anisotropic stiffness matrix $\mathbb{C}_{ijkl}$was obtained as follows, for each aortic segment:

$$\mathbb{C}_{ijkl} = \delta_{ik}\hat{t}^0{}_{lj} + \hat{t}^0{}_{il}\delta_{jk} + 4F^0_{iA}F^0_{jB}F^0_{kP}F^0_{lQ}\left.\frac{\partial^2 W}{\partial C_{AB}\partial C_{PQ}}\right|_{C^0}.$$

$F^0$, $C^0$, and $\hat{t}^0$ are the deformation gradient from an undeformed state, right Cauchy-Green tensor, and deformation-dependent Cauchy stresses:

$$F^0 = \begin{bmatrix} \frac{1}{\lambda_z^m \lambda_\theta^m} & 0 & 0 \\ 0 & \lambda_\theta^m & 0 \\ 0 & 0 & \lambda_z^m \end{bmatrix}$$

$$C^0 = (F^0)^T(F^0)$$

$$\hat{t}^0 = 2(F^0)\frac{\partial W}{\partial C^0}(F^0)^T$$

where $\lambda_z^m$ and $\lambda_\theta^m$ are the axial and the circumferential stretches, respectively, and $W$ is the strain energy function.

*Appendix C: Anisotropic material response estimation for electrowritten graft*

A finite element simulation was conducted to estimate the anisotropic material response of the electrowritten graft. Abaqus/Standard 2021 (Dassault Systemes, Providence, RI, USA) with static implicit solver was used for the simulation. The simulation aimed to replicate a displacement-controlled biaxial tensile test of 10x10-mm squared sample until a deformation of $\varepsilon_\theta = \varepsilon_z = 0.3$ (i.e., $\lambda_\theta = \lambda_z = 1.3$). The sample was deformed to 30% strain to replicate standard equibiaxial testing conditions (Tremblay et al.).

Due to the symmetrical shape of the sample, only a 5x5 mm patch was modeled and meshed with four shell elements (S4 elements in Abaqus library) with a 0.6-mm thickness. A two-fiber-family Holzapfel-Gasser-Ogden material model was used to simulate the mechanical response of the graft. The parameters were estimated by fitting the uniaxial data of Amato et al., resulting in the following parameters: $c_{10} = {}^{c}/_{2} = 0.005\ MPa,\ k_1 = 0.0143259\ MPa,\ k_2 = 2c_2 = 2.701\ MPa,\ \alpha = 50°$, $k = {}^{1}/_{3}$ (uniform distribution of fiber orientation), $D = 0$ (incompressible material). Figure 2 shows the set of boundary conditions that were applied to the patch. On the symmetry edges of the patch (i.e., AB and DA) symmetry conditions were applied (i.e., $u_z = M_z = M_\theta = 0$ on AB and $u_\theta = M_z = M_t = 0$ on DA) and displacements of $u_\theta = u_z = 1.5\ mm$ were applied on BC and CD, respectively. The simulation generated a plane stress field across the patch, and we obtained the curves that define the material's behavior in the axial and circumferential direction: axial Cauchy stress ($\sigma_z$) versus axial strain ($\lambda_z$) and circumferential Cauchy stress ($\sigma_\theta$) versus circumferential strain ($\lambda_\theta$).

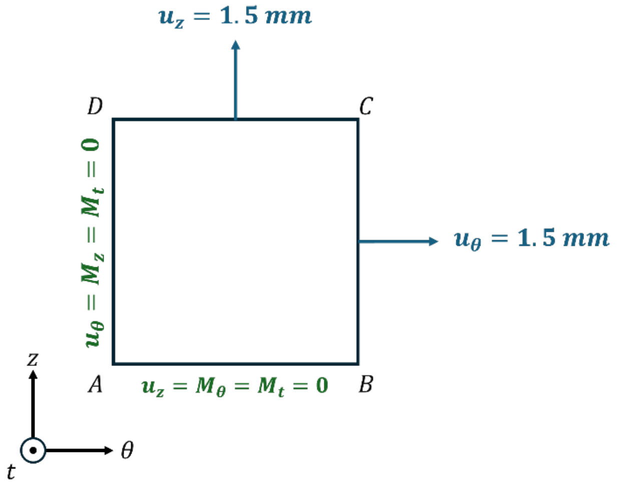


Figure 2: Set boundary conditions applied for the finite element simulation of the biaxial tensile test of the electrowritten material.
